# Distributions of Recorded Pain in Mental Health Records: A Natural Language Processing Based Study

**DOI:** 10.1101/2023.09.15.23295064

**Authors:** Jaya Chaturvedi, Robert Stewart, Mark Ashworth, Angus Roberts

## Abstract

**Objective:** The objective of this study is to determine demographic and diagnostic distributions of physical pain recorded in the clinical notes of a mental health electronic health records database by utilising natural language processing and to examine the level of overlap in recorded physical pain between primary and secondary care.

**Design, Setting and Participants:** The data were extracted from an anonymised version of the electronic health records from a large mental community and secondary healthcare provider serving a catchment of 1.3M residents in south London. These included patients under active referral and aged 18+ at the index date of July 1, 2018, and had at least one clinical document (>=30 characters) associated with their record between July 1, 2017 and July 1, 2019. This cohort was compared to linked primary care records from one of the four catchment boroughs.

**Outcome:** The primary outcome of interest was the presence or absence of recorded physical pain within the clinical notes of the patients. This does not include mental, psychological or metaphorical pain.

**Results:** A total of 27,211 patients were retrieved based on the extraction criteria. Of these, 52% (14,202) had narrative text containing relevant mentions of physical pain. Patients who were older (OR 1.17, 95% CI 1.15-1.19), female (OR 1.42, 95% CI 1.35-1.49), of Asian (OR 1.30, 95% CI 1.16-1.45) or Black (OR 1.49, 95% CI 1.40-1.59) ethnicities, and living in deprived neighbourhoods (OR 1.64, 95% CI 1.55-1.73) showed higher odds of recorded pain. Patients with an SMI diagnosis were found to be less likely to report pain (OR 0.43, 95% CI 0.41-0.46, p<0.001). When comparing the overlap between primary and secondary care, 17% of the CRIS cohort also had records within LDN, and 31% of these had recorded pain in both records.

**Conclusion:** The findings of this study show the sociodemographic and diagnostic differences in recorded pain, and have significant implications for the assessment and management of physical pain in patients with mental health disorders.

**Strengths and Limitations of this study:**

- This study utilises natural language processing on clinical notes to access a large sample with information about pain.
- This is the first cross-sectional study to summarise and describe the distribution of recorded pain within the clinical notes of mental health records.
- The recorded mentions of pain within clinical notes clearly depend on the patient sharing and the clinician recording their experiences.
- The findings are not generalisable to the general population since this study only looks at patients receiving mental healthcare within a specific geographic catchment.

## Introduction

### Background Rationale

Pain and its relationship with mental health are important research topics. Pain has imposed a significant burden on society in terms of medical care costs as well as lost productivity [1]. Pain is multifaceted, with physical, psychological, social, and biological causes and consequences [2]. Mental health disorders also present a considerable and complex public health problem, being a leading cause of disability and accounting for 28% of the national disease burden in the UK [3]. Electronic health records (EHRs) for mental health are a significant source of information for studying the intersection between pain and mental health within those who receive specialist service input. EHRs open the possibility of investigating how pain is recorded and its impact on clinical outcomes.

Severe mental illnesses (SMIs) include diagnoses of schizophrenia spectrum disorder, bipolar disorder, or severe major depressive disorder [4], where functional and occupational activities are severely impaired due to associated debilitating psychological problems [5]. While several studies have looked at the relationship between pain and schizophrenia and bipolar disorders [6–9] and at other mental illnesses such as depression [1,10–13], the complex and potentially bidirectional nature of this relationship requires further understanding. Analysis of secondary data sources, such as EHR databases, might help by providing a fuller picture of the recorded clinical presentation of this group of patients; however, a prerequisite is that pain is adequately represented in derived data.

Demographic features such as age, gender and ethnicity can influence pain perception and experiences. Pain affects twice as many persons over the age of 60 as it does younger individuals [14]. While pain is not a natural feature of the ageing process, many health conditions causing pain become more common with increasing age. Nonetheless, older patients often believe pain to be a normal aspect of ageing and might be hesitant when reporting it [14]. There have also been variations found in the reported perception of pain by female and male patients, with female patients reporting experiencing more pain than males [15,16]. Research has also shown disparities in pain perception across different ethnicities, with individuals of Black (African) ethnicity reporting greater pain than White counterparts [17].

Socioeconomic status (SES) plays a role in health and overall well-being, with deprivation associated with unfavourable health outcomes and increased mortality rates [18]. Patients with SMI already experience higher mortality rates than the general population, and this discrepancy is exacerbated by socioeconomic deprivation, primarily due to unequal access to good quality physical healthcare services [19–22]. Furthermore, patients with SMI continue to experience a decline in their SES over time, compounding its impact [23].

Most patient information is recorded in unstructured clinical narratives within EHR databases [24], and pain is likely to be no different, with few, if any, structured checklists ascertaining its presence in routine clinical care. Natural language processing (NLP), a computational approach to understanding and analysing human language, is therefore potentially useful for extracting such pain information. NLP has been applied extensively to EHR data, including studies of SMI, such as antipsychotic polypharmacy in mental health care [25], multimorbidity in individuals with schizophrenia and bipolar disorders [26], and extracting symptoms of SMI [27].

In addition to secondary care data, it is also useful to consider the recording of pain in primary care data. Within the UK, primary care is generally the first point of contact for patients [28]. Exploring the overlap of recorded pain between primary and secondary care could, therefore, provide a more comprehensive view of the patient’s pain experiences, and any discrepancies could highlight gaps in care and communication.

### Objectives

The objective of this study is to describe the distributions of recorded pain amongst mental health service users according to demographic factors such as age, gender and ethnicity, as well as neighbourhood deprivation levels and mental health diagnoses. This was achieved by examining recorded pain through the means of an NLP application within the clinical text of a mental health EHR database, and further evaluating this by measuring the overlap between pain recorded in secondary and primary health care, enabled through data linkage between the two.

## Methods

### Reporting

We use the RECORD [29] guidelines and checklist, an extension of the STROBE [30] guidelines, for reporting the results of this study.

### Setting

Data on recorded pain were obtained from the clinical text of a mental health EHR database, the Clinical Record Interactive Search (CRIS) resource. This contains a de-identified version of EHR data from The South London and Maudsley NHS Foundation Trust (SLaM), one of Europe’s largest mental healthcare organisations [31], which serves a geographic catchment of around 1.3 million residents in four south London boroughs (Croydon, Lambeth, Lewisham, Southwark). CRIS contains about 30 million free text documents, averaging 90 documents per patient [24].

Data were also obtained from a primary care database called Lambeth DataNet (LDN) [32], which accesses all GP records from general practices based in the London borough of Lambeth. Data linkages (at the patient level) are already in place between CRIS and LDN [33].

#### Ethical Approval

CRIS (as well as its associated linkages) has received ethical approval as a data resource for secondary analysis from the Oxford C Research Ethics Committee (reference 23/SC/0257). A patient-led oversight committee (detailed in [34]) reviews and approves research projects that use the CRIS database. For service users, an opt-out system is in place and is advertised in all promotional materials and campaigns. Only authorised individuals can access this data from within a secure firewall. The CRIS project approval references for this work are 21-021 and 23-003.

LDN approval was obtained as part of an existing CRIS project (project number 23-124) which included access to linked data from LDN (Caldicott Guardian approval, 15/9/21). This CRIS-LDN project aimed to examine the profile of patients with mental illnesses and chronic/persistent pain and compare them to controls from LDN who had chronic/persistent pain only.

#### Patient and Public Involvement

Patient and public involvement (PPI) in research is an active collaboration between researchers and members of the public, where the latter actively participate in contributing to the research [35]. A PPI group with lived experiences of SMI and chronic pain were consulted as part of this research. The nature of the data available was described to the group, and they were asked about their priorities regarding what research questions they would like answered. In response to this, the group was unanimously interested in further study of the differences in pain experiences based on demographics and diagnoses, and this was the main motivation for the objective of the study described here.

### Participants

A cohort of patients was extracted from the CRIS database comprising those who were active (i.e., under an accepted referral) and aged 18+ on the index date of July 1, 2018, and whose record contained at least one document (>= 30 characters) within a window of July 1, 2017 to July 1, 2019.

LDN extraction followed similar criteria for patients who were active on the index date, aged 18+, and contained pain diagnoses or medications from July 1, 2017 to July 1, 2019. Free-text information is unavailable within LDN, so no document criteria were required.

### Variables

#### Demographics

Age, gender, and ethnicity variables were extracted from structured tables within the CRIS database. Individuals with missing ethnicity values were retained as a separate category (Not stated/known).

#### Diagnosis

The primary diagnosis recorded closest to the index date of July 1, 2018, was extracted from the structured tables within the CRIS database. These are coded using ICD-10 [36]. The diagnosis codes were categorised as SMI (severe mental illnesses) and non-SMI, where SMI includes ICD-10 codes of F20-29 and F30-33.

#### Deprivation

The Index of Multiple Deprivation (IMD) decile measures from 2019 [37] were extracted for information on neighbourhood deprivation for each patient, based on their address at the time of the index date aggregated by Lower Super Output Area (LSOA) - a standard national administrative unit containing an average 1500 residents. National Census data are used to calculate IMD scores for each LSOA. A lower IMD decile indicates higher deprivation levels. Individuals with missing IMD scores were retained in a separate category.

#### Recorded Pain

Pain-related keywords generated from a lexicon of pain terms [38] were used to identify patients in the cohort who had mentions of physical pain recorded in their clinical notes within the predetermined window. An NLP application was used on the documents of these patients. The application classified sentences within the document as relevant or not, where relevant refers to a mention of physical pain affecting the patient, and not relevant refers to no or negated mentions, hypothetical mentions, and metaphorical mentions of pain. Only relevant mentions were used in the results reported here. The application has been described in detail in [39].

As with all other UK research based on access to anonymised primary care records, LDN does not allow access to any free text clinical notes. For this reason, pain information can only be extracted from the structured fields of the records. Read codes [40] were used to identify patients who had a pain diagnosis or were on any pain medications and treatments:

1. Pain medications code list - developed as part of a project described in [41], which focused on analgesics (obtained from dm+d (a dictionary of medicines and devices [42]) used in the treatment of 35 long-term conditions. These 35 conditions were obtained from [43], a cross-sectional study on multimorbidities in patients registered with 314 medical practices in Scotland as of March 2007.
2. Pain diagnosis and treatments code list - developed as part of a collaboration project with Outcomes Based Healthcare (OBH), an organisation that provides a platform for the study of population health outcomes [44], with the research described in [45].

While these codes were developed for chronic pain, they are generic enough to be used for this research. These code lists are available on GitHub^1^.

##### Anatomy Related to Recorded Pain

Another NLP application was developed for identifying anatomy mentioned in relation to pain. This was a classifier that generated a binary output - “mentioned” or “not mentioned”. This application was run on sentences labelled as relevant by the pain application. Once the sentences that contained mentions of body parts were identified, they were run through MedCATTrainer [46], which used named entity recognition (NER), a type of NLP task to label entities within the text to identify the specific body parts mentioned within the text. The purpose of using MedCATTrainer was that it linked the identified body parts to unique identification numbers (SCTID) from SNOMED CT, a terminology of clinical terms. These SCTIDs were used to aggregate the mentioned body parts, for ease of analysis. For example, foot, calf, and knee mentions would be aggregated under “lower limb”.

##### Overlap between CRIS and LDN

To examine the overlap across primary (LDN) and secondary (CRIS) care, the patient IDs from the CRIS cohort (N=27,211) were searched for matching records within the LDN database over the same window of July 1, 2017 to July 1, 2019. Variables were generated indicating the presence of the patients within LDN, along with variables indicating the presence of any codes for pain medication, diagnosis or treatment based on the predefined lists described above. This allowed the identification of patients with documented pain experiences in both their mental health and primary care records for the aligned time period. The cross-referencing process enabled the comparison of recorded pain between the two systems at the patient level.

### Descriptive Statistics

All analysis was conducted using STATA v15.1 and the Python programming language (version 3.10.0).

Descriptive statistics were obtained for demographic, deprivation and diagnosis features and compared between the two groups - patients who had recorded pain (class 1) and those who did not (class 0) - within their clinical notes. Chi-squared tests and logistic regression were conducted between the two classes to obtain adjusted odds ratios. Frequencies of body parts affected by pain and the overlap of recorded pain experiences between CRIS and LDN were also reported.

## Results

### Data Extraction

Based on the extraction criteria, 27,211 patients were represented. Amongst these patients, 18,188 had pain keywords mentioned within their documents. These documents were run through the NLP application to label them as relevant to pain (class 1) or not (class 0), resulting in 14,202 patients who had relevant mentions of pain within their clinical notes (Figure 1).

**Figure 1.**
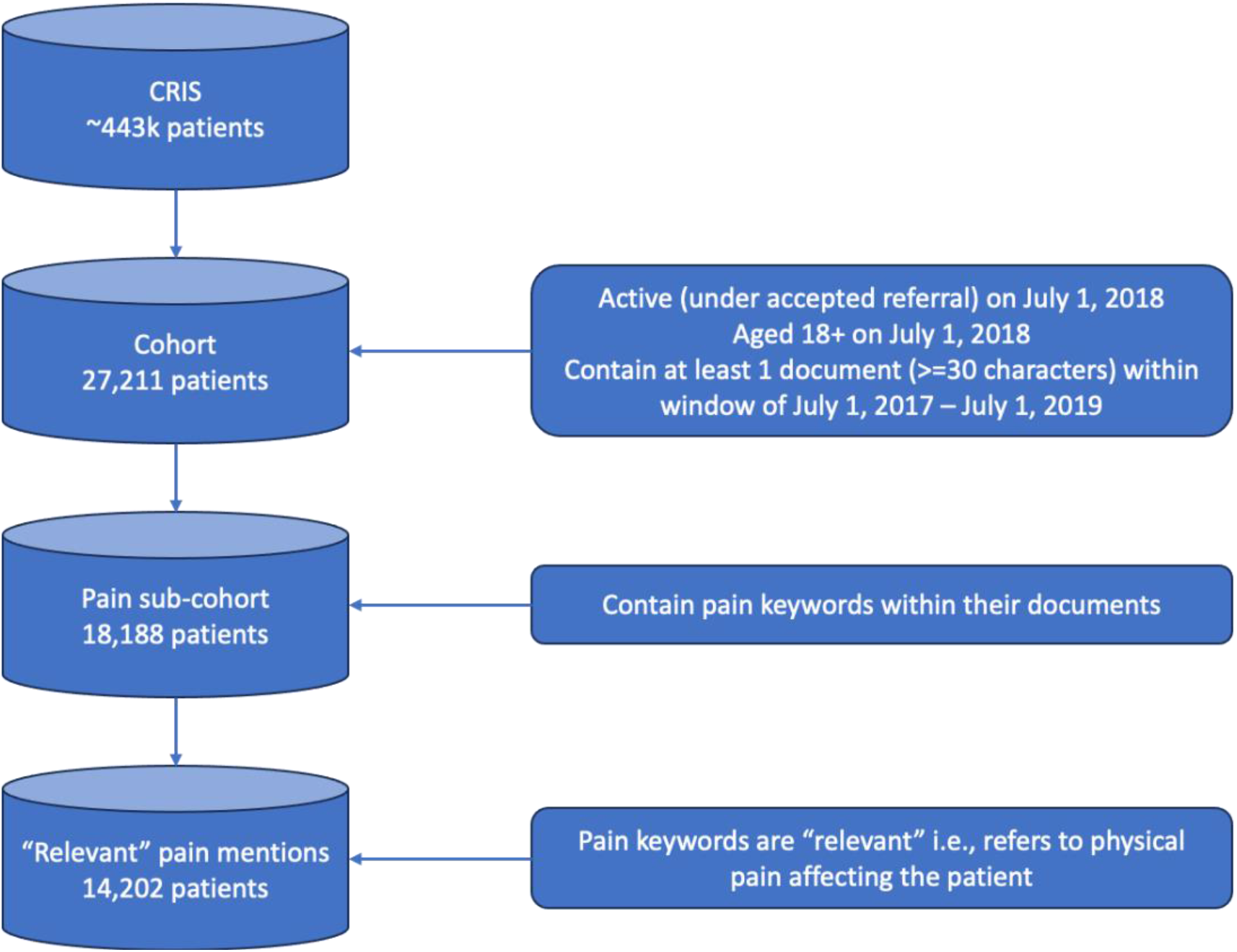
Data Extraction.

### Cohort Characteristics

Amongst the cohort of 27,211 patients, the mean age of the cohort was 44 (Inter-quartile range 29-55, SD 17.5), with 50.3% female and 48.2% of White ethnicity. The majority of the cohort (72.2%) lived in more deprived areas (IMD score <=5), and 67.0% received a non-SMI diagnosis. 66.8% of the patients (18,188 patients and 174,167 mentions within documents) contained pain keywords within their documents, and 52.1% of the cohort (14,202 patients) contained relevant mentions of pain in their documents.

### Pain Mentions

Records of 52.1% of the patients within the cohort contained relevant mentions of pain. Differences between the patients who showed recorded pain (class 1) in their clinical notes and those who didn’t (class 0) are shown in Table 1. Class 0 includes patients who did not have any pain mentions in their documents, as well as patients whose pain mentions were classified as not relevant. Patients within class 1 had an average of 10 pain mentions within their documents.

**Table 1.** Distributions between the two classes - class 0 (no recorded pain) and class 1 (recorded pain)

| Characteristic | n | Class 0<br>(no recorded pain) | Class 1<br>(recorded pain) |
| --- | --- | --- | --- |
| N (%) | 27,211 | 13,009 (47.9) | 14,202 (52.1) |
| Mean Age<br>(IQR) | 44<br>(29–55) | 41<br>(27–52) | 46<br>(32–56) |
| Gender (N, %) |  |  |  |
| Male | 13,471 | 7,037 (54.1) | 6,434 (45.3) |
| Female | 13,709 | 5,953 (45.7) | 7,756 (54.6) |
| Not known | 31 | 19 (0.2) | 12 (0.1) |
| Ethnicity (N, %) |  |  |  |
| White | 13,139 | 6,014 (46.2) | 7,125 (50.1) |
| Black | 5,866 | 2,115 (16.2) | 3,751 (26.4) |
| Not stated/known | 4,708 | 3,418 (26.2) | 1,290 (9.0) |
| Asian | 1,506 | 592 (4.5) | 914 (6.4) |
| Other | 1,197 | 512 (3.9) | 685 (4.8) |
| Mixed | 795 | 358 (2.7) | 437 (3.0) |
| Index of Multiple Deprivation (N, %)<br>Decile 2019 |  |  |  |
| <= 5 (more deprived) | 19,660 | 8,847 (68.0) | 10,813 (76.1) |
| > 5 (less deprived) | 6,686 | 3,836 (29.4) | 2,850 (20.0) |
| Not known | 865 | 326 (2.5) | 539 (3.9) |
| Primary Diagnosis: SMI vs Non-SMI<br>(ICD-9 code) (N, %) |  |  |  |
| SMI | 8,962 | 3,059 (23.5) | 5,903 (41.5) |
| Non-SMI | 18,249 | 9,950 (76.5) | 8,299 (58.5) |

Demographic variations emerged between those with/without recorded pain in the cohort, as shown in Table 1. The mean age was higher in patients with recorded pain at 46 (SD=17) compared to 41 (SD=17) for the remainder. Patients with recorded pain were more likely to be female and had a higher representation across all ethnic minorities. Additionally, patients with documented pain experiences were more likely to live in higher deprivation neighbourhoods.

Table 2 presents demographic, deprivation and diagnostic associations with recorded pain obtained through logistic regressions (unadjusted and adjusted for different factors as detailed below).

**Table 2.** Logistic Regression findings for variables reflecting differences in class 0 (no recorded pain) and class 1 (recorded pain) (N = 27,211) Values are given as odds ratio (95% CI), and * indicates significance at *p* < 0.001 Model 1 contained the demographic variables only [age, gender and ethnicity]. Model 2 contained the variables from Model 1, plus the variable for deprivation (IMD Decile]. Model 3 contained the variables from Model 2 plus the diagnosis variable. Model 4 contains the ethnicity and deprivation variables alone.

|  | Logistic Regression models |  |  |  |  |
| --- | --- | --- | --- | --- | --- |
|  | Unadjusted | Mutually adjusted |  |  |  |
|  |  | Model 1 | Model 2 | Model 3 | Model 4 |
| **Age** (per 10 years) | 1.17<br>[1.15, 1.19] * | 1.12<br>[1.11, 1.14] * | 1.12<br>[1.11, 1.14] * | 1.11<br>[1.10, 1.13] * | - |
| **Gender** |  |  |  |  |  |
| Male | 1 (reference) | 1 (reference) | 1 (reference) | 1 (reference) | - |
| Female | 1.42<br>[1.35, 1.49] * | 1.42<br>[1.35, 1.49] * | 1.43<br>[1.36, 1.50] * | 1.47<br>[1.40, 1.55] * | - |
| Not known | 0.69<br>[0.33, 1.42] | 1.08<br>[0.50, 2.33] | 1.06<br>[0.49, 2.30] | 1.10<br>[0.51, 2.38] | - |
| **Ethnicity** |  |  |  |  |  |
| White | 1 (reference) | 1 (reference) | 1 (reference) | 1 (reference) | 1 (reference) |
| Asian | 1.30<br>[1.16, 1.45] * | 1.36<br>[1.22, 1.52] * | 1.34<br>[1.19, 1.49] * | 1.21<br>[1.08, 1.36] * | 1.29<br>[1.15, 1.44] * |
| Black | 1.49<br>[1.40, 1.59] * | 1.58<br>[1.48, 1.69] * | 1.50<br>[1.40, 1.60] * | 1.25<br>[1.17, 1.34] | 1.42<br>[1.33, 1.52] * |
| Other | 1.12<br>[1.00, 1.27] | 1.20<br>[1.06, 1.36] | 1.17<br>[1.03, 1.32] | 1.10<br>[0.97, 1.24] | 1.08<br>[0.96, 1.33] |
| Mixed | 1.03<br>[0.89, 1.18] | 1.15<br>[0.99, 1.33] | 1.12<br>[0.96, 1.30] | 1.06<br>[0.91, 1.23] | 1.01<br>[0.87, 1.17] |
| Not known | 0.31<br>[0.29, 0.34] * | 0.36<br>[0.34, 0.39] * | 0.37<br>[0.34, 0.40] * | 0.40<br>[0.37, 0.44] * | 0.32<br>[0.30, 0.35] * |
| **Index of Multiple Deprivation** |  |  |  |  |  |
| National Decile <=5 | 1.64<br>[1.55, 1.73] * | - | 1.43<br>[1.35, 1.51] * | 1.37<br>[1.29, 1.45] * | 1.41<br>[1.33, 1.50] * |
| **Diagnosis** |  |  |  |  |  |
| SMI | 0.43<br>[0.41, 0.46] * | - | - | 0.56<br>[0.53, 0.59] * | - |

Unadjusted odds ratios revealed patients with documented pain experiences were more likely to be older (OR 1.17, 95% CI 1.15-1.19, p<0.001), female (OR 1.42, 95% CI 1.35-1.49, p<0.001), of Asian (OR 1.30 in relation to a White reference group, 95% CI 1.16-1.45, p<0.001) or Black (OR 1.49, 95% CI 1.40-1.59, p<0.001) ethnicities, and living in deprived neighbourhoods (OR 1.64, 95% CI 1.55-1.73, p<0.001) when compared to the remainder of the sample. In a model containing all demographic variables (Model 1), the odds ratios were strengthened for all ethnic minority groups. Additional adjustment for neighbourhood deprivation (Model 2) resulted in a further strengthening of the odds ratio for females. In the model also adjusted for diagnoses (Model 3), odds ratios became stronger for females. Patients with SMI had lower odds of documented pain (OR 0.43, 95% CI 0.41-0.46, p<0.001) than non-SMI patients, with the odds ratio slightly weakening when adjusted for demographics, deprivation and diagnosis (Model 3). A supplementary model (Model 4) including both ethnicity and deprivation as covariates showed independent increased odds for Asian and Black patients and those in more deprived neighbourhoods.

### Anatomy Distributions

Additional descriptive data were generated on the nature of the pain reported. Amongst the 14,202 patients with any recorded pain, there were 174,167 mentions of pain within the documents. Of these, 7,555 (53%) patients included 40,418 mentions of the anatomy associated with the pain. Of these 53%, each patient had an average of 5 body parts mentioned in the context of pain. The most common body part affected by pain, as per the recorded mentions, was lower limbs, which accounted for 20% of all mentions where anatomy could be ascertained (Table 3).

**Table 3.** Body parts affected (at mention level)

| Body Part | Mentions | Frequency<br>(mention-level) |
| --- | --- | --- |
| Lower limbs | Feet, ankle, leg, knee, calf, thigh, toes | 20% |
| Upper body, excluding back | Chest, side of chest, upper body, torso | 19% |
| Upper limbs | Hand, wrist, arm, elbow, thumb, shoulder | 17% |
| Stomach/abdomen region | Stomach, abdomen, groin, bladder, prostate | 16% |
| Head and neck | Head, tooth, face, mouth, tongue, eye, ear, neck | 15% |
| Non-specific site | Entire body, skin, muscle, joint | 8% |
| Back | Back, lower back | 5% |

### Overlap with Primary Care

When comparing secondary care CRIS records with those of primary care from LDN, among the 27,211 patients of the CRIS cohort, 4,822 patients (17%) also had records in LDN. Amongst these patients who had records in both CRIS and LDN, 1,507 (31%) patients were identified as having some recorded instance of pain in both their records, while 687 (14%) patients showed recorded pain only in LDN (primary care). Among the 27,211 patients within CRIS, 12,695 (46%) had recorded pain only within CRIS (mental health care), as seen in Figure 2.

**Figure 2.**
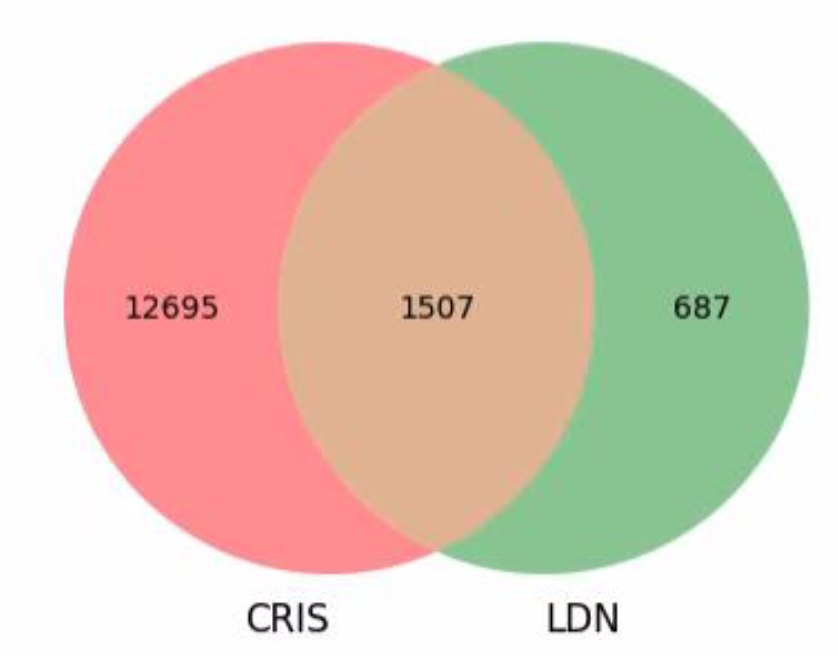
Overlap of recorded pain in CRIS and LDN.

## Discussion

This study investigated the differences observed in recorded pain mentions within the clinical notes of mental health records. The results reflect current literature findings that pain is a common issue among patients with mental health disorders. In a cohort of 27,211 patients, 18,188 (67%) patients contained pain-related keywords in their text, and 14,202 (52%) patients had relevant pain mentions, i.e., the mention indicated physical pain affecting the patient in question. Disparities in recorded pain mentions were found across genders, with females being over-represented. This is consistent with other research that indicates gender disparities in pain experiences [16,48,49]. Furthermore, while patients with known ethnicities were mostly over-represented in the cohort of relevant pain mentions (in relation to those with unknown ethnicity), most noticeable were the Black, Asian and other ethnic groups. This aligns with research around the undertreatment of patients within certain ethnic minority groups [50] and highlights the need for a comprehensive exploration of pain experiences across diverse populations. Moreover, the study’s findings are also consistent with studies that indicate the impact of deprivation on health outcomes [18], as people living in more deprived areas (IMD decile <= 5) were more frequently recorded with pain.

When comparing the overlap of patients between primary and secondary care, it was found that 17% of the patients within the CRIS cohort also had records within LDN. Amongst these patients, 31% had recorded pain instances in both records. While this overlap between primary and secondary care seems low, it is important to bear in mind that Lambeth only represents 22% of the catchment covered by CRIS [47]. Patients present in CRIS but not in LDN could include patients who have recorded instances of pain within the free-text clinical notes in LDN and might have been missed in this study since we do not have access to this text. Furthermore, this study did not differentiate between acute and chronic pain mentions and focused on extracting mentions of physical pain of any duration. As a result, the higher occurrence of pain mentioned within CRIS can be partially attributed to the documentation of such acute or short-lived pain episodes. Conversely, the GP records within LDN likely focus on recording persistent and chronic pain experiences. This disparity in recording pain should be considered when interpreting the findings of this study. Looking specifically at chronic pain instances within the CRIS notes may improve the comparability. However, the temporal information required to determine pain chronicity from clinical notes is a particular challenge and can be difficult to extract reliably. Future work can attempt to differentiate acute and chronic pain through temporal or contextual information, which could provide richer insight. However, the current broad inclusion of pain provides wider coverage for this initial exploration of pain mentioned within clinical notes.

A strength of this study is the size of the data set available and the access to information about pain from the clinical text. To the best of our knowledge, this is potentially the first cross-sectional study to summarise and describe the distribution of recorded pain derived from routine mental health records. While the cohort data extraction did not apply any filters on demographics, aiming for broad representativeness, other systemic biases related to access to healthcare resources may still exist. Factors like deprivation level and ethnicity can influence the utilisation of services and, therefore, documentation within health records, often stemming from perceived barriers to access. However, by not restricting cohort selection on demographic factors, this study intended to capture a diverse patient population receiving care across the South London boroughs.

A limitation of this study is that the recorded mentions of pain within clinical notes depend on the clinician recording them. The actual occurrences of pain experiences could remain unaccounted for if they weren’t recorded by the clinicians or were not shared with the clinicians, especially for patients with severe mental illnesses who might be completely or partially nonverbal. While the NLP application achieved good performance metrics during its development and evaluation, it is not impervious to imperfections. Instances of pain experiences might have been overlooked if they were not included as examples during the training of the application.

The scope of this study is limited to the examination of mental health records from an EHR database in South London. Given the absence of a comparative cohort of patients experiencing pain without any mental health disorders, the findings of this study are not generalisable to the overall population. However, they might be relevant and generalisable to some extent to other populations of patients with mental health disorders. It is essential to acknowledge the potential influence of gender and ethnicity on the reporting of pain experiences, particularly if females and minority ethnicities (due to language barriers or other reasons) are less likely to self-report their pain experiences [50,52,53]. Since the focus of this study has been on a mental health EHR database, the clinical care within this setting is focused on mental health issues reported by the patients. Consequently, as much importance might not be given to the investigation and reporting of physical health conditions such as pain.

This study cannot determine a cause-and-effect relationship or directionality between pain and mental illnesses. Despite this, the study has highlighted existing disparities in recorded pain experiences and brings to attention the need for further research to better understand and address them at the point of care.

## Conclusion

The outcomes of this study have significant implications for the assessment and management of pain amongst patients with mental health disorders and highlight the importance of utilising NLP methods on EHR databases for research purposes. Notably, these findings reiterate the recommendations set forth by Mental Health America [54], advocating the need for proactive initiation of conversations around mental health and pain with patients. Relying solely on patients to self-report symptoms could potentially lead to worse outcomes, especially since the stigma surrounding pain and mental health conditions may prevent patients from seeking the necessary treatment. Thus, early and proactive interventions could go a long way towards improved long-term outcomes. Unfortunately, there still exists a perceived lack of credibility and empathy towards patients living with pain [55], particularly when compounded by co-existent mental illnesses. This was one of the main points shared by the PPI group consulted as part of this study. More research in this area can help towards these issues and provide safer and equitable access to good-quality pain management.

While these findings represent a step forward, they are only one side of the story. Combining these findings with patient-reported insights could offer a more comprehensive understanding of pain experiences within this cohort. However, achieving this is a challenging task due to the lack of such data and the inability to link patient-reported experiences to their health records. Further research is needed to better understand the relationship between pain and mental health and to develop more effective interventions to manage pain in this population.

## Supporting information

RECORD checklist

## Data Availability Statement

Data are owned by a third party, Maudsley Biomedical Research Centre (BRC) Clinical Records Interactive Search (CRIS) tool, which provides access to anonymised data derived from SLaM electronic medical records. These data can be accessed by permitted individuals from within a secure firewall (i.e. the data cannot be sent elsewhere) in the same manner as the authors. For more information, please contact. Any STATA and Python code used in this project will be available on GitHub^2^.

## Author Contributions

The idea was conceived by JC, AR, and RS. JC conducted the analyses and drafted the manuscript. MA provided insights on LDN data. AR and RS provided guidance in the design and interpretation of results. All authors commented on drafts of the manuscript and approved the final version.

## Funding and Acknowledgements

AR is funded by Health Data Research UK, an initiative funded by UK Research and Innovation, Department of Health and Social Care (England) and the devolved administrations, and leading medical research charities. RS is part-funded by i) the National Institute for Health Research (NIHR) Biomedical Research Centre at South London and Maudsley NHS Foundation Trust and King’s College London; ii) the National Institute for Health Research (NIHR) Applied Research Collaboration South London (NIHR ARC South London) at King’s College Hospital NHS Foundation Trust; iii) the DATAMIND HDR UK Mental Health Data Hub (MRC grant MR/W014386); iv) the UK Prevention Research Partnership (Violence, Health and Society; MR-VO49879/1), an initiative funded by UK Research and Innovation Councils, the Department of Health and Social Care (England) and the UK devolved administrations, and leading health research charities. JC is supported by the KCL-funded Centre for Doctoral Training (CDT) in Data-Driven Health. The funders were not involved in the study design, collection, analysis, interpretation of data, the writing of this article or the decision to submit it for publication. RS declared research support received in the last 36 months from Janssen, GSK and Takeda. All other authors declare no other competing interests.

This paper represents independent research part-funded by the National Institute for Health Research (NIHR) Biomedical Research Centre at South London and Maudsley NHS Foundation Trust and King’s College London. The views expressed are those of the authors and not necessarily those of the NHS, the NIHR or the Department of Health and Social Care. The funders had no role in study design, data collection and analysis, decision to publish, or preparation of the manuscript.

This work uses data provided by patients and collected by the NHS as part of their care and support. An application for access to the Clinical Record Interactive Search (CRIS) database for this project was submitted and approved by the CRIS Oversight Committee. The authors would like to acknowledge Dr Ruimin Ma for her help in obtaining the LDN codes.

## Footnotes

1 https://github.com/jayachaturvedi/pain_in_mental_health

2 https://github.com/jayachaturvedi/pain_in_mental_health

